# Feasibility and Performance of Procalcitonin-guided antimicrobial stewardship during autologous stem cell transplantation

**DOI:** 10.64898/2025.12.15.25340973

**Authors:** Anupam Pande, Sandra Susanibar Adaniya, William Clark, Rachel Wilkinson, Monica Grazziutti, Senu Apewokin

## Abstract

**Background:** Antibiotic stewardship during stem cell transplantation (SCT) is challenging.. Procalcitonin (PCT) has been employed successfully in critical care patients to safely guide stewardship. However, procalcitonin guided stewardship has not been robustly assessed in SCT recipients. We sought to evaluate the potential utility of PCT to guide antimicrobial de-escalation during engraftment.

**Methods:** 100 SCT patients were prospectively enrolled in a “strategy trial” and had infectious complications documented. Lab parameters - CBC, BMP, CRP were obtained daily as standard of care (SOC) while PCT was obtained for research purposes. Providers were blinded to PCT results. We compared duration of antimicrobial escalation between actual events (SOC model) and a proposed PCT model. In this hypothetical PCT model, antibiotic de-escalation would occur if CRP remained <100 mg/dl and PCT <0.25 ng/ml after 3 days of escalation. Escalation events were defined as a substitution or addition of an antimicrobial agent after initiation of prophylactic antimicrobials.

**Results:** 77 patients had escalation events and of these, 33 had bacterial infections. A total of 136 antimicrobial escalations events were identified, and of these only 39(28.7%) were associated with documented infections. The standard of care model had a mean duration (+SD) of 9.08 (+ 6.08) antibiotic days. If the PCT model were employed, the mean duration (+SD) would be 4.44 (+ 6.16) days (p<0.001). The PCT model, however, would have missed 11 infections\

**Conclusion:** Procalcitonin-guided antimicrobial stewardship during autologous stem cell transplantation is feasible however optimization is necessitated for utilization as a tool to guide antibiotic prophylaxis during SCT.

## Background

Infectious complications still account for a significant proportion of morbidity and mortality events associated with stem cell transplantation. ^1^ Infectious disease management in stem cell transplant (SCT) recipients is particularly challenging because transplant recipients often do not mount typical immune responses that characterize an immunocompetent host defense to infectious pathogens. ^2^ As a result, clinical and physiologic parameters are frequently unreliable or often absent. Until recently, clinical diagnostic methods have been limited by low sensitivity and an inability to identify disease processes in a timely manner. Recently, biomarkers of infectious processes have been developed to enable clinicians improve diagnostic capabilities and achieve better outcomes. One such biomarker, procalcitonin (PCT), is often elevated in patients with bacterial and fungal infections, but not elevated in viral infections.^3^ Procalcitonin has been utilized in stem cell recipients to provide discrimination between uninfected and infected patients with bacterial and/or fungal organisms.^4,5^ It can also be utilized as an indicator of the overall prognosis of patients.^6^ Unfortunately, previous studies have examined the utility of procalcitonin as a stand-alone diagnostic test for patient prognosis.^6^ While useful for examining patient prognosis, they shed minimal insight on feasibility of procalcitonin as an adjunct for other diagnostic tests discriminating infectious processes, as well as procalcitonin’s utility for guiding antimicrobial stewardship. Furthermore, lack of blinding has made it impossible to control for provider biases, and the value of serial biomarker testing is not often assessed.^7^ Surveillance for some viral and fungal pathogens using assays such as CMV PCR (cytomegalovirus polymerase chain reaction), Beta-D-Glucan (BDG) and Aspergillus Antigen (serum galactomannan) is increasingly becoming common practice in hematopoietic stem cell Transplant patients. ^8,9^ However, bacteria-specific surveillance is still not widely performed, likely due to the absence of a robust assay. We postulated that if used appropriately, procalcitonin could potentially serve as surveillance for bacterial infections and thereby impact the utilization of antimicrobial agents. The purpose of this study is to evaluate potential utility of serial procalcitonin measurements to guide antimicrobial de-escalation during engraftment employing a rigorous study design.

### Methods

A “strategy trial” (Figure 1) was performed on patients being treated for Multiple Myeloma or lymphoma with high dose chemotherapy and stem cell support from the day of stem cell infusion until the day of engraftment, or 30 days post-transplant (which ever occurred first). To do this, 100 patients undergoing autologous SCT were prospectively enrolled, and all infectious complications documented. Lab parameters including complete blood count (CBC), basic metabolic panel (BMP) and C-reactive protein (CRP) were obtained daily from the day of transplant until engraftment as part of standard of care. For research purposes only, PCT was measured on aliquots obtained for CRP measurements. Serum galactomannan, BDG and CMV PCR were also performed three times a week as standard of care for all patients. Clinicians were blinded to PCT results. We compared duration of antimicrobial escalation between actual events (standard of care model) and the hypothetical scenario where treating clinicians de-escalated antibiotics based upon prespecified triggers (PCT model). Escalation events were defined as the substitution or addition of an antimicrobial agent after initiation of prophylactic antimicrobials. Prophylactic antimicrobials were initiated at the onset of chemotherapy. Infections were documented by microbiological, radiological, or serological data. In this hypothetical PCT model, antimicrobial de-escalation intervention point occurred if there was an absence of documented infection, CRP of <100 mg/dl, and PCT <0.25 ng/ml, within three days of an escalation event.^10^,^3^ We also compared the performance of procalcitonin surveillance as a discriminator between infected and uninfected patients during the engraftment admission. Additionally, we estimated the utility of serial procalcitonin values measured up to fourteen days after transplantation by constructing receiver operator curves (ROC) that compared serial white blood cell count (WBC), CRP and a combination of WBC, PCT and CRP. Finally, we determined the optimal cutpoints that discriminate between infected and uninfected patients for each of the four parameters (WBC, CRP, PCT, temperature) by determining J-index (also known as Youden’s index).

**Figure 1.**
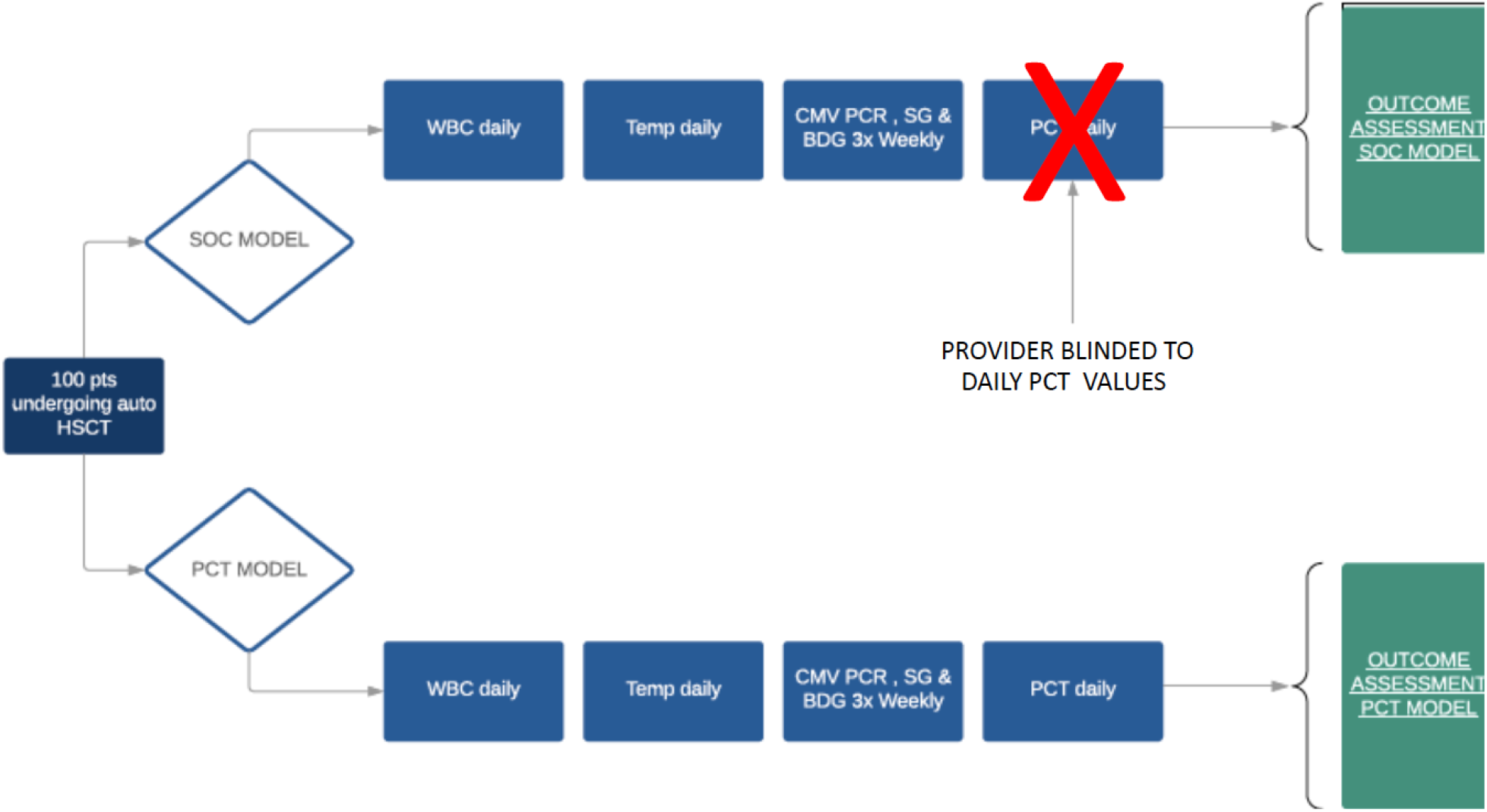
Depiction of surveillance assays for standard of care and procalcitonin models

## Results

Infections were documented in 41 of 100 patients enrolled. These patients had 51 total infections and of these, 33 were bacterial, with 16 being blood stream infections (BSIs). Twenty percent of the total infections in the cohort (10 of 51) were clinically documented or radiologically diagnosed infections (i.e., cellulitis or pneumonia on chest computed tomography with negative respiratory panel or respiratory cultures) and 80.3 percent (41 out of 51) were microbiologically confirmed infections. Majority of these infections occurred at an average of 8.5 (SD +/-4.8) days after SCT (Table 1). The mean WBC at the time of diagnosis was 0.92. Escalation events were documented among 77 patients, with a total of 136 events. The total days of antimicrobial escalation was 9.08 (SD +/-6.08) using the standard of care (SOC) model. The most utilized antibiotics during escalation days were cefepime in 32% of patients, ertapenem in 30%, and vancomycin in 28% of patients. Implementation of the PCT model would have reduced the mean number of escalation days from 9.08 (SD +/-6.08) to 4.44 (SD +/-6.16) (Figure 2). Using PCT as a discriminator of documented bacterial infections, an area under the curve (AUC) of 10.31 (SD +/-5.29) was achieved among the infected vs 6.10 (SD +/-2.99) in the uninfected (Figure 3). The corresponding AUCs for CRP was 951.5 (SD +/-133.1) among the infected and 725.5 (SD +/-121.9) in the uninfected (figure 4). Fever as a discriminator achieved an AUC of 1274 (SD +/-2.39) for infected 1274 (SD +/-2.67) in uninfected patients, respectively. The mean temperature on day of infection diagnoses was 98.5 degrees Fahrenheit. Similarly, the corresponding PCT and CRP were 0.98 and 54.3 respectively. The performance of these biomarker parameters as depicted by ROC curves are seen in Figure 5; PCT was 70.2%, temperature was 67.6%, WBC was 66.4% and CRP was 64.0%. The J-index (Youden’s index) was 0.32 ng/ml, 98.4 degrees Fahrenheit, 1.11 and 29.2 mg/L for PCT, Temp, WBC and CRP respectively.

**Table 1.**
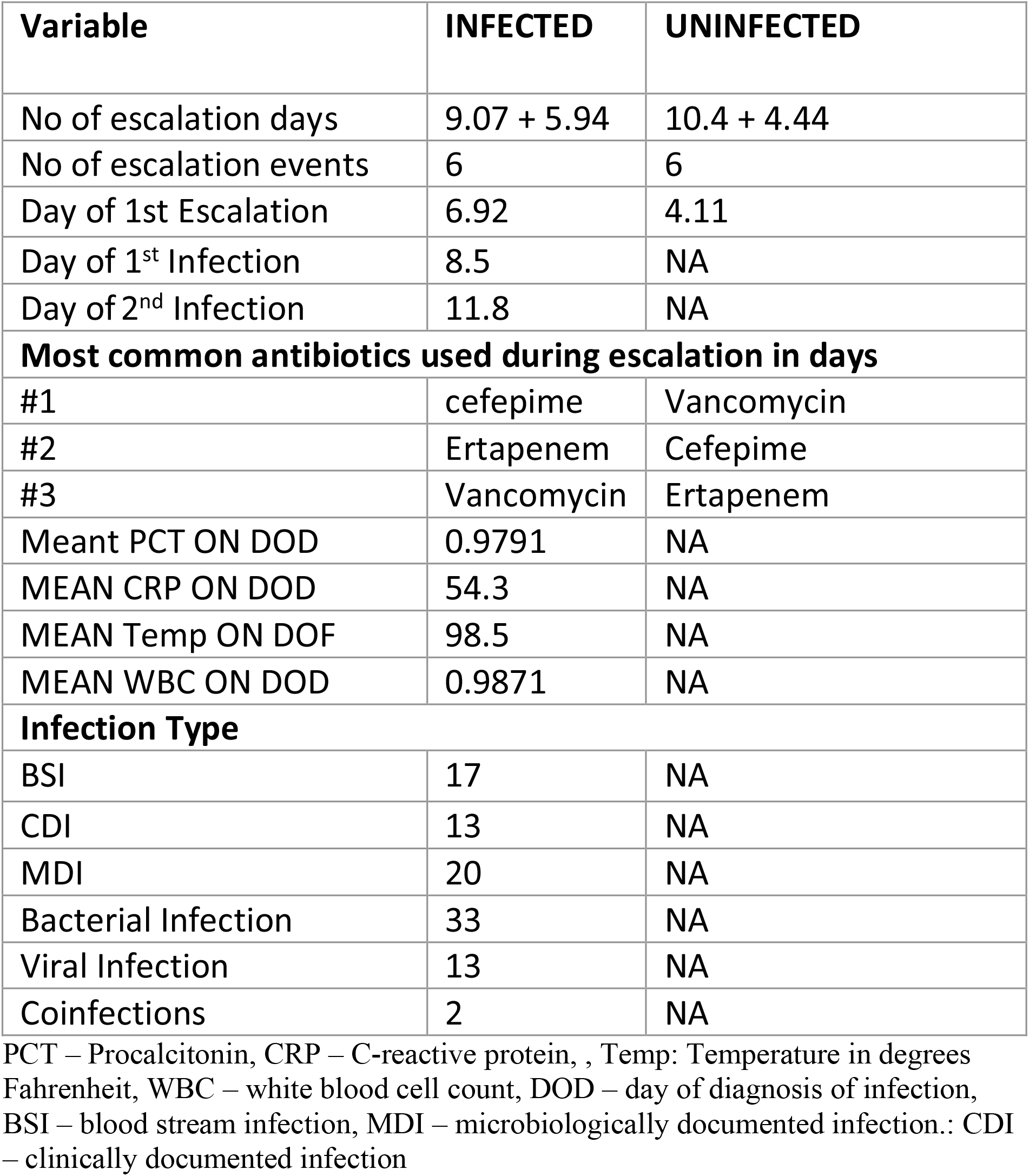
Comparison of antibiotic-related events and infections between infected and uninfected patients.

**Figure 2.**
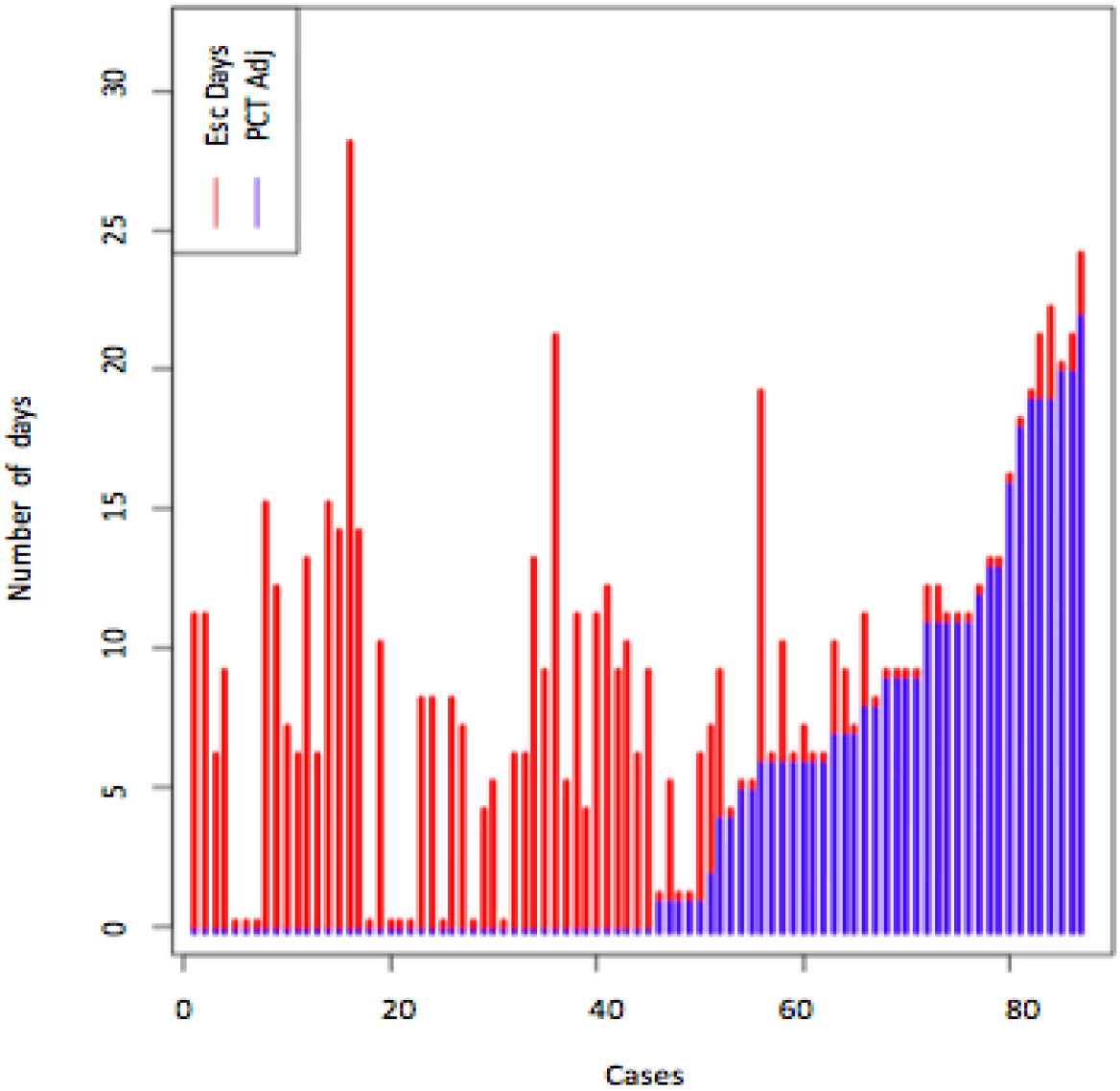
Comparison of antibiotics escalation days between standard of care model and procalcitonin-adjusted models.

**Figure 3.**
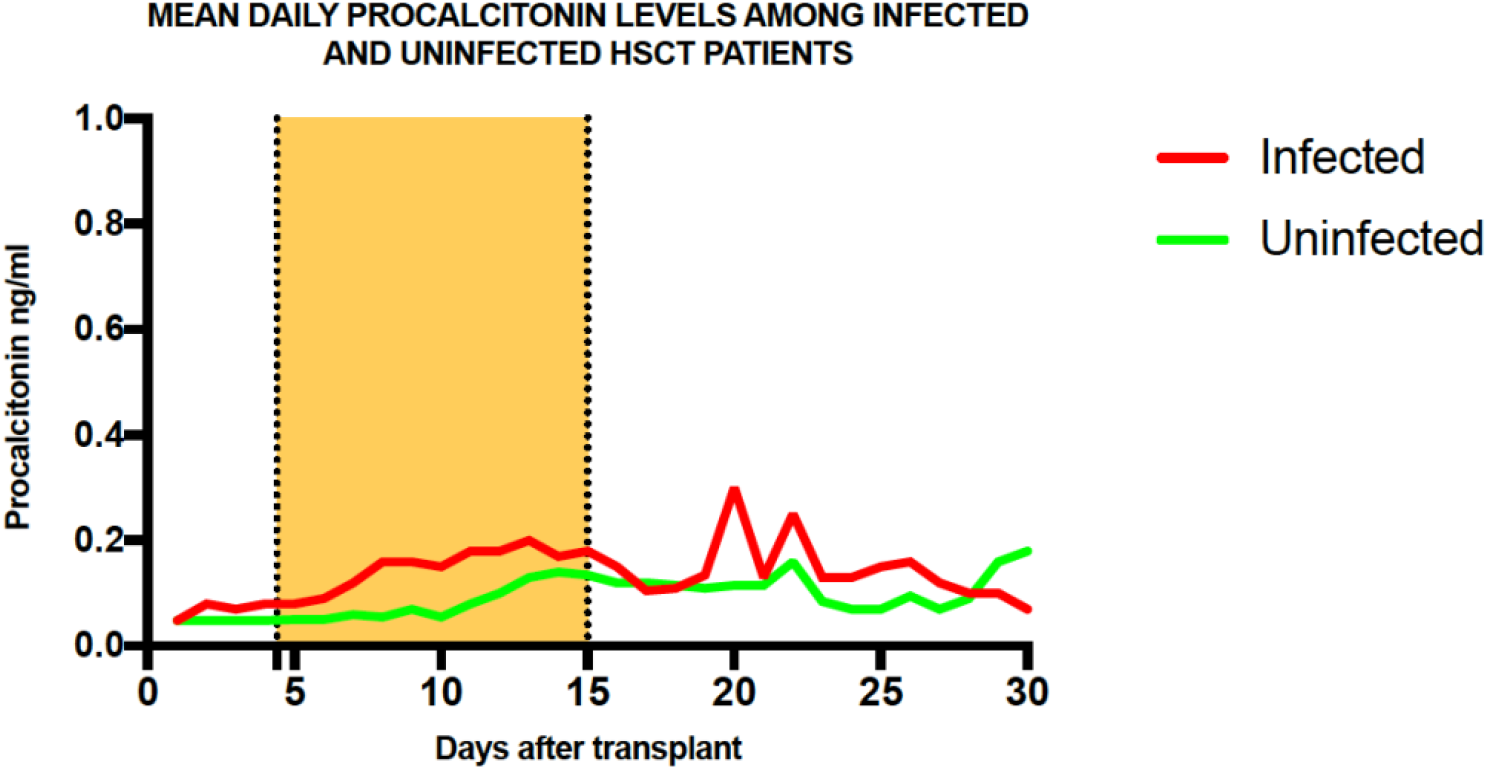

**Figure 4.**
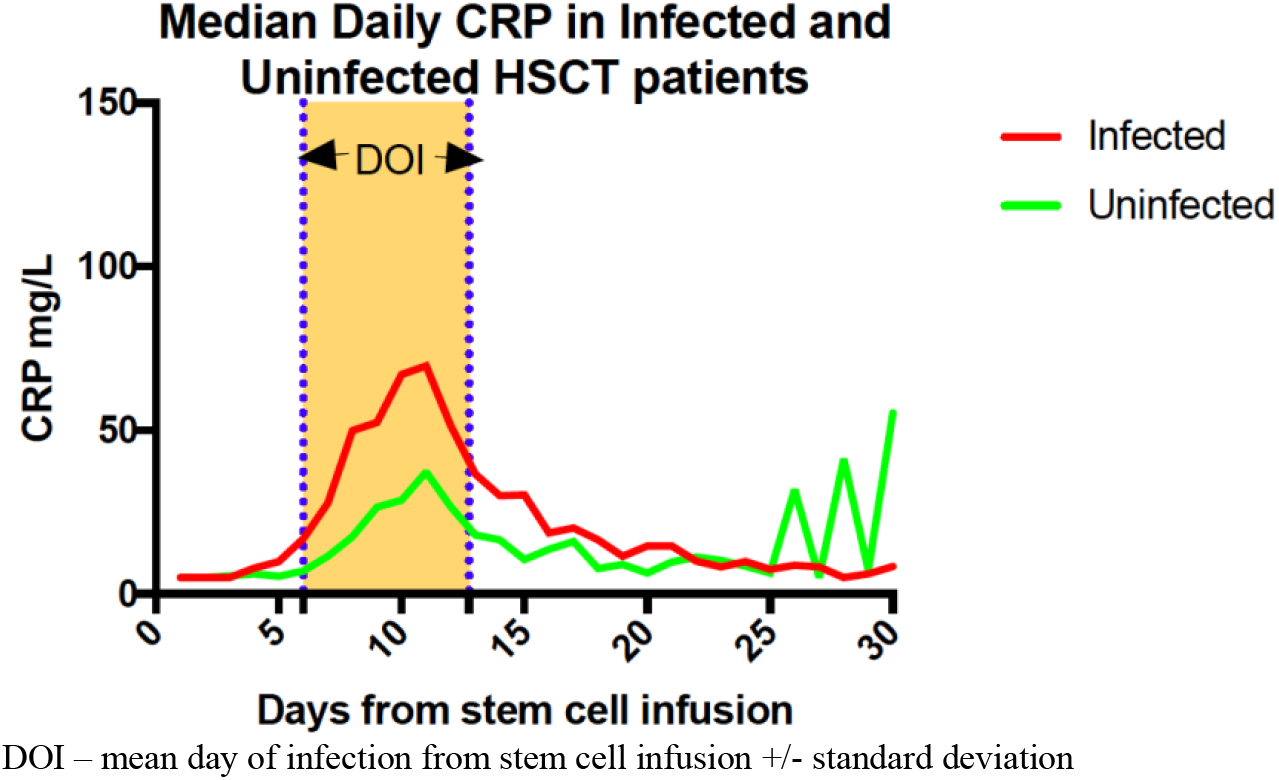

**Figure 5.**
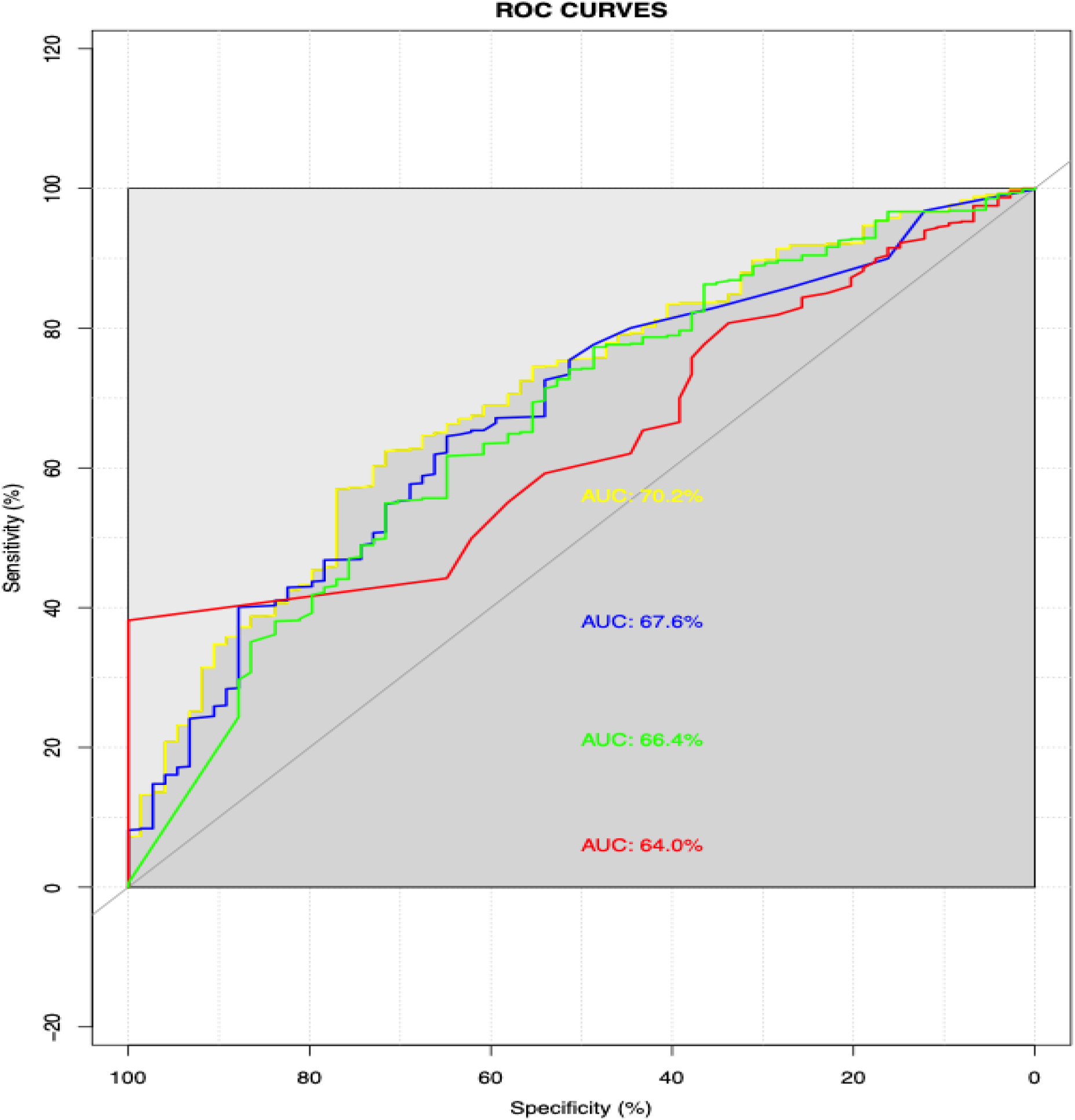
ROC curves (PCT – yellow; Temp – blue ; WBC – green; CRP – red)

## Discussion

The use of antimicrobial prophylaxis has become conventional clinical practice, and is incorporated into the National Comprehensive Cancer Network (NCCN) guidelines for prevention of infection in cancer patients.^11^ The evidence driving these practices was provided by several landmark studies conducted in the early 2000s.^12,13^ Over the last few years, rapid advancements have been made in diagnostic microbiology, and the incorporation of rapid diagnostic tests into clinical care has markedly reduced the time to initiation of appropriate antibiotics. ^14^ Biomarker surveillance guided preemptive strategies, coupled with rapid diagnostics may make the case for withholding prophylaxis, and employing biomarker strategies to guide antimicrobial utilization. Unfortunately, such “strategy trials” are sparse and thus evidence to evaluate the efficacy of biomarker approaches are limited. We herein report a strategy trial employing procalcitonin surveillance to guide antimicrobial use in SCT recipients. Strengths of this study include the prospective design, provider blinding, and the use of serial biomarker measurements rather than single measurement reported in existing literature. Other unique features of this study include the use of procalcitonin in conjunction with other biomarkers and diagnostics rather than as a stand-alone test. Another highlight is that we provide optimal cutpoints for PCT in the population studied which hitherto was unclear. It is notable that even with current infection control and prevention strategies, the incidence of infections in SCT patients is still high. A previous study evaluating complications following autologous SCT in multiple myeloma patients found a 45.7% rate of complications within the first 100 days following transplantation, and among these patients, 79.9% encountered an infectious complication.^15^ Overall, this study found an infection prevalence of 36.5% in patients within 100 days of autologous stem cell transplantation, consistent with infection rates in our study.

A prior study by Homsi et al evaluated infections in cancer patients admitted to a palliative care unit, in which infection prevalence was 40%, consistent with infection rates in our study. Homsi et al also noted that 15-20% of these infections were polymicrobial. ^16^ Comparatively, in our study, 25 % of all infections were associated with coinfections, and 12 % of study subjects were coinfected with viral pathogens. Given such high incidence of infections in these patients surveillance strategies with PCT has been proposed as potential solution given the ability to distinguish bacterial and fungal infections from viral infections.^17^

Escalations events occurred in 77 percent of subjects in the study cohort. This indicates the vast majority patients in the cohort were exposed to potentially avoidable, but ultimately inappropriate antibiotic regimens as only 33% of them displayed bacterial infections; in other words, for each patient who had an appropriate antibiotic intervention, two others received inappropriate antibiotic intervention. A model that could reduce inappropriate use by even fifty percent will go a long way towards aiding antimicrobial stewardship efforts. The proposed PCT models reduced days of antibiotic exposure by fifty percent, reflected by a reduction in both the number of escalation events as well as the duration of escalations. Utilization of this model could potentially impact the development of multiple drug resistant organisms (MDROs), as the most common antibiotics used during escalations were broad-spectrum agents. The studies that inspired the use of near-universal prophylactic antibiotic intervention in SCT recipients were conducted in the early 2000s, where rapid diagnostics and culture independent methods for diagnosing infections were not in use. Culture independent molecular assays have transformed clinical microbiology and thus value of universal prophylaxis in the current era needs to be revisited. ^18^

### Performance of fever, WBC, CRP and PCT as discriminators for bacterial infections 48hrs prior to diagnosis

#### Fever

Fever is often the trigger for antibiotic escalation in neutropenic patents. Providers placed standing orders that trigger initiation of antipyretics for temperatures above 100.3 degrees Fahrenheit. It was therefore impossible to assess the performance of fever as a discriminator in this study.

#### CRP

Our study findings that CRP was able to distinguish infected patients from non-infected are consistent with other studies. Fassas et al have reported that a CRP velocity associated of around 15-24hrs is strongly associated with infections, and our findings corroborate and report a similar increase.^3^ CRP is hampered by decreased specificity, but does have reasonable sensitivity if steroids and/or chemotherapy are not concurrently being administered. Provided that most of the infections occurred after chemotherapy infusions were complete, the sensitivity limitation will not be overly present in this patient group.

#### PCT

PCT was seemingly equal to CRP in discriminating between bacterially infected and non-infected patients. However, PCT can augment CRP during receipt of chemotherapy and when the use of steroids and other agents could reduce the performance of CRP. Use of such agents could also suppress fevers, hence delaying the diagnosis of infections. Another use for PCT could be to screen out patients who are poor candidates for SCT, such as those with uncontrolled infection. While this study did not utilize PCT to screen out poor candidates for SCT, this approach should be considered in future studies, as noted on the experience of patient #2 in our study. The patient suffered a cardiac arrest on Day 2 post SCT and was found to have E. *coli* bacteremia post-mortem. Upon unblinding, we noted that this patient had a PCT of 40 and a CRP of 9.4 on the day of SCT. We postulate that PCT screening prior to SCT may have prompted further work up and potentially delayed SCT until the reason for elevation was resolved. A potential explanation for the rather low CRP is that the patient was on active chemotherapy at the time they developed the bacteremia. Many studies have evaluated PCT in cancer patients, with varying sensitivity and specificity, dependent on the relative PCT cutoff employed. The negative predictive value (NPV) is considered a vital index in such patients given that these patients may have antibiotic intervention withheld in the event of a false negative PCT and could ultimately see dire consequences as a result. The advantage of utilizing CRP as an adjunct test is that it partly aids in overcoming this limitation. Additionally, the use of other diagnostic modalities such as cultures and radiologic studies could potentially identify any missed cases. In our study, the sensitivity (73% of procalcitonin was similar to that reported and specificity of 63.3%). It is possible that viral coinfections would contribute to a higher rate of false-negative procalcitonin values. It is notable that 2(6%) of the bacterial infections had viral coinfections. Viral infections have been reported to increase interferon gamma levels, which in turn suppresses PCT levels, leading to a falsely low or even negative PCT level in patients with bacterial and viral coinfections. ^19^

## Limitations

PCT measurement began on the day of SCT, thus the impact of chemotherapy infusion on levels could not be assessed. The PCT model missed 11 infections and thus its utility as a stand-alone test to detect infectious processes will not be ideal. However, this can be mitigated by the utilization of other diagnostic measures, such as cultures, that will still be performed in the setting of fevers irrespective of a negative PCT, thus identifying infections missed by PCT monitoring. PCT use in de-escalation will not result in lack of protection. PCT has been noted to be particularly sensitive for patients with systemic inflammatory response syndrome (SIRS), or sepsis, therefore making it less likely to remain negative, even in the setting of missed infection, should that infection continue to progress.^20^ False negatives could have been due to viral coinfections; however, this could be alleviated if interferon gamma levels are ascertained with the negative sample to ensure PCT levels are not artificially suppressed.

## Future directions

The use of bacterial prophylaxis has become part of mainstream practice and incorporated in the NCCN guidelines. The evidence inspiring these practices were provided by several landmark studies conducted in the early 1990s. Over the last few years, several biomarkers have been evaluated to guide antimicrobial stewardship in infectious diseases. Biomarker surveillance guided preemptive strategies coupled with rapid diagnostics make the case for withholding prophylaxis and employing preemptive strategies if robust biomarkers or biomarker panels could be developed.

## Conclusions

Our study provides evidence that while the use of PCT to guided antibiotics use is feasible, false negative tests may occur, and further optimization is warranted to improve its performance in SCT patients.

## Data Availability

All data produced in the present work are contained in the manuscript

## Notes

### Competing Interest Statement

The authors have declared no competing interest.

### Clinical Trial

NCT02035189

### Funding Statement

This study did not receive any funding

### Author Declarations

University of Arkansas Institutional Review Board

